# PSMA PET SUVmax Adds Information Beyond Conventional Risk Indices in Localized Prostate Cancer: A Prospective Lesion-Level Radiopathologic Study

**DOI:** 10.64898/2026.09.04.26362267

**Authors:** Georges (Camille) Motchoffo Simo, Pranav Movva, Jiyoun Seo, Bohang Shou, Daniel Vega, Theodore Hagens, Evan C. Norris, Daniel J. Luthringer, Alessandro D’Agnolo, Kyung Hyun Sung, Jeremie Calais, Wayne G. Brisbane, Robert E. Reiter, Timothy J. Daskivich, Debiao Li, Rola Saouaf, Hyung L. Kim, Adam B. Weiner

**Author notes:** **Corresponding Author:** Georges (Camille) Motchoffo Simo, Department of Urology, Cedars-Sinai Medical Center, 8635 W. Third Street, Suite 1070W, Los Angeles, CA 90048.

## Abstract

**Background:** Prostate-specific membrane antigen (PSMA) positron emission tomography (PET) is increasingly used to characterize newly diagnosed prostate cancer on the assumption that uptake highlights aggressive disease. However, maximum standardized uptake value (SUVmax) varies widely across localized tumors. Recent genomic assessments have suggested SUVmax reflects tumor biology even when accounting for other tumor indices. Here, we examined highly annotated pathologic, imaging and clinical correlates of intraprostatic SUVmax at the level of individual tumor foci in a prospective multimodal cohort.

**Methods:** Exploratory lesion-level analysis of the radical prostatectomy arm of a single-center prospective diagnostic-accuracy trial (NCT04461509). Patients underwent simultaneous ^18^F-DCFPyL PSMA PET, multiparametric magnetic resonance imaging (MRI) and high-resolution MRI before prostatectomy, with central blinded pathology and imaging review. Grade Group (GG) ≥2 foci measuring ≥10 mm were eligible. The primary outcome was focus-level SUVmax. Correlates spanning histology, tumor microenvironment, MRI and clinical indices were examined by Spearman correlation and rank-based group comparison, followed by separate linear regressions of log(1+SUVmax) each adjusted for GG and tumor size. A digital-pathology sensitivity analysis quantified immune, stromal and tumor pixel-area fractions on hematoxylin and eosin (H&E) sections from a subset of foci using a trained QuPath classifier.

**Results:** Twenty-seven patients contributed 34 tumor foci, of which 21 (61.8%) were GG2. Median SUVmax was 5.7 (interquartile range [IQR] 3.9–12.1; range 0.0–57.9). SUVmax rose across the cribriform/intraductal carcinoma (Crib/IDC) gradient (median 3.9, 4.8 and 10.2 for absent, focal and extensive; p = 0.010), correlated with % Gleason pattern 4 (ρ = +0.47, p = 0.005) and correlated inversely with stromal content (ρ = −0.40, p = 0.020). Grade Group (p = 0.115), Prostate Imaging Reporting and Data System (PI-RADS) 4 versus 5 (p = 0.705), standard apparent diffusion coefficient (ADC) (ρ = −0.27, p = 0.130), baseline prostate-specific antigen (PSA) (ρ = +0.04, p = 0.816) and PSA density (ρ = −0.02, p = 0.918) were not associated with SUVmax. In separate models of log(1+SUVmax) adjusted for Grade Group and tumor size, % Gleason pattern 4 remained associated with uptake (standardized β = +0.76, 95% CI +0.11 to +1.41), Crib/IDC (β = +0.36, −0.03 to +0.74), and stromal content (β = −0.30, −0.62 to +0.02). On 18 slides re-quantified by pixel classifier, no pixel-based compartment fraction correlated with SUVmax (immune ρ = −0.06, stroma ρ = +0.02, tumor ρ = −0.04; all p > 0.5).

**Conclusions:** In this prospective lesion-level cohort, intraprostatic PSMA SUVmax was associated with increased Gleason pattern 4 and cribriform/intraductal burden but not with other common risk indices. SUVmax therefore appears to carry information additive to conventional risk factors. Further investigations examining the tumor biology that reflects SUVmax are warranted.

## Introduction

Prostate-specific membrane antigen (PSMA) positron emission tomography (PET) has become a standard imaging modality for staging intermediate- and high-risk prostate cancer and for selecting patients for PSMA-targeted radioligand therapy.^1–3^ In newly diagnosed disease its use rests on the assumption that PSMA expression can detect all or most clinically aggressive cancer. In practice, uptake in treatment-naïve localized tumors is heterogeneous, and as many as one in five localized International Society of Urological Pathology (ISUP) Grade Group (GG) ≥2 tumors show low or absent uptake.^4,5^ What that heterogeneity represents, and how much of it is already reflected by the indices clinicians use to stratify risk, remains incompletely defined.

Radiology–pathology correlation studies have generally reported ISUP GG as the principal pathologic correlate of the maximum standardized uptake value (SUVmax).^6,7^ Higher grade is associated on average with higher SUVmax, but within-grade variance is substantial and grade alone does not account for the high-grade tumors that show little uptake.^8^ Cribriform and intraductal carcinoma (Crib/IDC) are adverse-prognosis architectures that are biologically distinct from conventional Gleason pattern 4 and have been examined in relation to PSMA uptake,^9^ but have not been modeled as an ordinal exposure alongside grade in a prospectively accrued PSMA PET–pathology cohort. The tumor microenvironment, and stromal content in particular, is a further candidate source of variation that has rarely been quantified alongside imaging.^10^

Prior work from our group has shown PSMA expression in treatment-naïve localized prostate cancer reflects underlying tumor biology, including susceptibility to androgen deprivation and radiotherapy.^8^ Whether that biology is already captured by the pathologic, imaging and clinical metrics used in routine risk stratification is unknown. In that context, we asked which histologic, microenvironmental, MRI and clinical features are associated with intraprostatic SUVmax at the level of individual tumor foci, in a prospective cohort with highly annotated data from simultaneous ^18^F-DCFPyL PSMA PET, multiparametric magnetic resonance imaging (mpMRI) and high-resolution MRI (hrMRI) before radical prostatectomy (RP).

## Materials and Methods

### Study design and participants

This is an exploratory, lesion-level radiology–pathology correlation analysis of the radical-prostatectomy arm of a single-center prospective diagnostic-accuracy trial (NCT04461509) conducted at Cedars-Sinai Medical Center between April 2021 and September 2024. The parent trial compared ^18^F-DCFPyL PSMA PET combined with hrMRI against standard mpMRI for intraprostatic cancer detection. Patients subsequently underwent either RP or focal high-intensity focused ultrasound. Only the RP arm contributed to the present analysis. No patient received neoadjuvant treatment. This analysis was approved by the Cedars-Sinai Institutional Review Board (STUDY00003844) and all participants provided written informed consent under the trial protocol. Reporting follows the Strengthening the Reporting of Observational Studies in Epidemiology (STROBE) guideline. Full parent-trial eligibility criteria and the pre-specified analysis plan are given in **Supplemental Methods**. Imaging acquisition parameters are tabulated in **Table S1**.

Of 62 patients enrolled in the parent trial, 33 were assigned to the RP arm (**Figure S1**). Only foci that were GG ≥2 and at least 10 mm in greatest dimension on central pathology review were analyzed, to limit the influence of partial-volume effects and tumor size on SUVmax. Six patients had no focus meeting both criteria and were excluded, leaving 27 patients.

### Imaging

Imaging was reviewed centrally for this analysis. MRI was interpreted by a dedicated genitourinary radiologist (E.N.) blinded to pathology, who scored each lesion by the Prostate Imaging Reporting and Data System (PI-RADS) version 2.1.^11^ PET was interpreted separately by a nuclear-medicine physician (A.D.) blinded to the mpMRI read and to pathologic grade. Each pathology-defined focus was matched to a volume of interest on the fused PET/MRI images using the prostate cylindrical coordinate system,^12^ and SUVmax was recorded as the primary PET metric.

Reference-tissue SUVs (blood pool, bone marrow, parotid) and the secondary PET metrics are defined in **Supplemental Methods** and summarized in **Table S2**. Foci in which no voxel within the matched volume of interest exceeded surrounding benign prostatic background were recorded as SUVmax = 0; because a standardized uptake value of exactly zero is not physically attainable in perfused tissue, these denote uptake indistinguishable from adjacent benign prostate rather than absence of tracer signal, and are interpreted throughout as no measurable uptake above background.

### Pathology

Each specimen was reviewed and annotated by a genitourinary pathologist (D.J.L.). Tumor foci were defined as spatially contiguous carcinoma separated from neighboring foci by benign parenchyma, and multi-slice foci were checked for contiguity to avoid duplication. Foci were localized using the prostate cylindrical coordinate system previously validated at our institution (**Figure S2**).^12^ For each focus we recorded primary and secondary Gleason patterns, ISUP Grade Group, % Gleason pattern 4 and pattern 5, Crib/IDC status, extraprostatic extension (EPE), seminal vesicle invasion (SVI), and visually estimated stromal and immune-cell percentages. Pathologic nodal status was recorded per patient. Invasive cribriform and intraductal carcinoma were scored together as a single ordinal variable (absent, focal, extensive) because the two architectures frequently coexist within one focus and were not separately quantifiable in every focus. Extent was graded semiquantitatively according to the proportion of the focus occupied by cribriform and/or intraductal architecture across the reviewed sections, without a fixed area cutoff.

### Digital pathology

As a pre-specified sensitivity analysis to test whether the visually estimated microenvironment measures were reproducible by an independent method, a pixel classifier was trained in QuPath v0.7.0 and applied to separate H&E whole-slide sections from the same tumor blocks, quantifying immune, stromal and tumor compartments as pixel-area fractions. Agreement with the visual estimates was judged against a pre-specified primary criterion of an intraclass correlation of at least 0.60 for immune percentage, with a Spearman rank correlation of at least 0.50 as a secondary criterion. After quality control, 21 slides from 20 patients were analyzable; three had contributed training swatches and were excluded from the concordance analysis to keep it out of sample, leaving 18 slides from 17 patients, one of whom contributed two slides.

These were the foci for which a separate section had been cut, scanned and passed quality control, and are not a random sample of the 34 analyzed foci. Classifier training, region-of-interest annotation and quality-control criteria are described in **Supplemental Methods**, and per-slide values are given in **Table S3**. A representative classifier overlay is shown in **Figure S3**.

### Outcomes and covariates

The primary outcome was focus-level SUVmax as a continuous variable. Candidate correlates were pathologic (Grade Group, % Gleason pattern 4 and 5, Crib/IDC, EPE, SVI, nodal status), microenvironmental (stromal and immune percentage), size-related (greatest pathologic dimension, ellipsoid tumor volume), imaging (PI-RADS, prostate zone, T2-weighted signal intensity, and both standard and high-resolution apparent diffusion coefficient [ADC] and diffusion-weighted imaging [DWI] signal intensity) and clinical (baseline prostate-specific antigen [PSA], MRI prostate volume, PSA density). Secondary PET metrics (SUVmean, PSMA volume, PSMA-total, tumor-to-reference ratios) and a comparison of foci above and below a pre-specified SUVmax threshold of 4, adopted from the parent-trial protocol and from published data linking low SUVmax to low PSMA expression,^13^ are reported in **Supplemental Results**.

### Statistical analysis

Descriptive statistics are median (interquartile range [IQR]) for continuous variables and n (%) for categorical variables. Associations between SUVmax and continuous covariates were assessed by Spearman rank correlation; comparisons across categorical variables used the Kruskal–Wallis or Mann–Whitney test. This analysis is exploratory and hypothesis-generating: no adjustment for multiple comparisons was applied, no single association is presented as confirmatory, and p values are reported to convey the strength of individual associations and should be read in that light. For adjusted analyses we fit separate ordinary least-squares regressions of log(1+SUVmax), one per predictor of interest, each adjusted for ISUP Grade Group (ordinal factor: GG2, GG3, GG4–5) and tumor greatest dimension. Continuous predictors and the Crib/IDC ordinal score were standardized to one standard deviation (SD), so coefficients are expressed per 1 SD and are comparable across predictors. Because Grade Group is itself defined in part by the proportion of Gleason pattern 4, the pattern 4 coefficient estimates within-grade variation in pattern 4 burden rather than an effect independent of grade. Only seven of 27 patients contributed two foci and the within-patient intraclass correlation on the modeled log scale was <0.01, so models were fit without a clustering adjustment. Agreement between pixel-classifier and pathologist estimates was assessed by intraclass correlation (two-way, absolute agreement, single measures) and Bland–Altman analysis. Tests were two-sided with α = 0.05. All analyses are fully reproducible from a single R Markdown file, available in the repository cited in the **Data Availability Statement**.

## Results

### Cohort

Twenty-seven patients contributed 34 tumor foci. Seven patients (26%) contributed two foci each. Median age at surgery was 65.9 years (IQR 61.4–71.0), median baseline PSA 6.9 ng/mL (IQR 6.0–9.7), median MRI-derived prostate volume 41.4 mL (IQR 30.5–50.1) and median PSA density 0.19 ng/mL/mL (IQR 0.10–0.31). Most patients were White (67%) or Asian (26%; **Table 1**).

**Table 1.** Clinicopathologic, imaging and PET characteristics.

| Characteristic | Value |
| --- | --- |
| **Patient characteristics (n = 27)** |  |
| Age at surgery, years | 65.9 [61.4, 71.0] |
| Baseline PSA, ng/mL | 6.9 [6.0, 9.7] |
| MRI prostate volume, mL | 41.4 [30.5, 50.1] |
| PSA density, ng/mL/mL | 0.19 [0.10, 0.31] |
| **Race** |  |
| White | 18 (66.7%) |
| Asian | 7 (25.9%) |
| Black | 1 (3.7%) |
| Other | 1 (3.7%) |
| **Clinical T stage** |  |
| T1c | 18 (66.7%) |
| T2a | 5 (18.5%) |
| T2b | 2 (7.4%) |
| T2c | 2 (7.4%) |
| Patients contributing two foci | 7 (25.9%) |
| **Pathologic features (n = 34 foci)** |  |
| **Grade Group** |  |
| GG 2 | 21 (61.8%) |
| GG 3 | 6 (17.6%) |
| GG 4 | 2 (5.9%) |
| GG 5 | 5 (14.7%) |
| % Gleason pattern 4 | 30 [15, 60] |
| % Gleason pattern 5 (n = 13 with pattern 5) | 10 [5, 10] |
| **Cribriform / intraductal carcinoma** |  |
| Extensive | 18 (52.9%) |
| Focal | 9 (26.5%) |
| Absent | 7 (20.6%) |
| **Extraprostatic extension** |  |
| None | 17 (50.0%) |
| Focal | 9 (26.5%) |
| Extensive | 8 (23.5%) |
| Seminal vesicle invasion | 7 (20.6%) |
| Pathologic nodal status (pN1) | 5 (14.7%) |
| **Tumor size (n = 34 foci)** |  |
| Greatest dimension, mm | 20.0 [15.0, 26.8] |
| Estimated tumor volume, cm<sup>3</sup> | 1.10 [0.60, 2.99] |
| **Tumor microenvironment (n = 34 foci)** |  |
| Immune cell % (n = 29 quantified) | 1.0 [1.0, 5.0] |
| Stromal % | 20.0 [20.0, 30.0] |
| **MRI (n = 34 foci)** |  |
| **Prostate zone** |  |
| Peripheral | 26 (76.5%) |
| Transition | 5 (14.7%) |
| Both | 3 (8.8%) |
| PI-RADS 4 (of 33 MRI-visible) | 17 (51.5%) |
| PI-RADS 5 (of 33 MRI-visible) | 16 (48.5%) |
| Standard ADC, $\times 10^{-6}$ mm<sup>2</sup>/s (n = 33) | 621 [512, 675] |
| Standard DWI signal intensity (n = 33) | 29 [27, 34] |
| T2W signal intensity (n = 33) | 129 [107, 151] |
| High-resolution ADC (n = 30) | 546.5 [477.5, 626.5] |
| High-resolution DWI signal intensity (n = 30) | 47.5 [42.0, 63.0] |
| **PSMA PET (n = 34 foci)** |  |
| SUVmax | 5.7 [3.9, 12.1] |
| SUVmean | 3.5 [2.4, 6.8] |
| SUVmax < 4 | 9 (26.5%) |

Continuous variables are median [IQR]; categorical variables are n (%). Patient-level variables are reported for 27 patients and focus-level variables for 34 tumor foci.
Immune cell percentage was quantified in 29 foci; the remaining 5 foci were recorded as <1% and were coded as 0.5 for analysis.
PI-RADS percentages are among the 33 MRI-visible foci; the remaining focus scored PI-RADS <3.
Abbreviations: ADC, apparent diffusion coefficient; DWI, diffusion-weighted imaging; GG, ISUP Grade Group; IQR, interquartile range; PI-RADS, Prostate Imaging Reporting and Data System; PSA, prostate-specific antigen; PSMA, prostate-specific membrane antigen; SUV, standardized uptake value; T2W, T2-weighted.

GG2 foci were most common (21 of 34, 61.8%). Crib/IDC architecture was extensive in 18 foci (52.9%), focal in 9 (26.5%) and absent in 7 (20.6). Median greatest dimension was 20.0 mm (IQR 15.0–26.8) and median estimated tumor volume 1.10 cm³ (IQR 0.60–2.99). Thirty-three foci were MRI-visible: 17 scored PI-RADS 4 and 16 scored PI-RADS 5. Median SUVmax across the 34 foci was 5.7 (IQR 3.9–12.1) with a range of 0.0 to 57.9.

### SUVmax associated with tumor architecture

Across the categorical features, Crib/IDC architecture showed a graded association with SUVmax. Median SUVmax was 3.9 in foci without cribriform or intraductal architecture, 4.8 where it was focal, and 10.2 where it was extensive (Kruskal–Wallis p = 0.010; **Figure 1A**). SUVmax did not differ by GG (p = 0.115; **Figure 1B**). Median SUVmax was 5.1 in GG2 (n = 21), 12.1 in GG3 (n = 6), 21.4 in GG4 (n = 2) and 5.0 in GG5 (n = 5). EPE (p = 0.100), SVI (p = 0.701), PI-RADS 4 versus 5 (p = 0.705) and prostate zone (p = 0.826) were not associated with SUVmax (**Figure 1**, **Table 2**).

**Figure 1.**
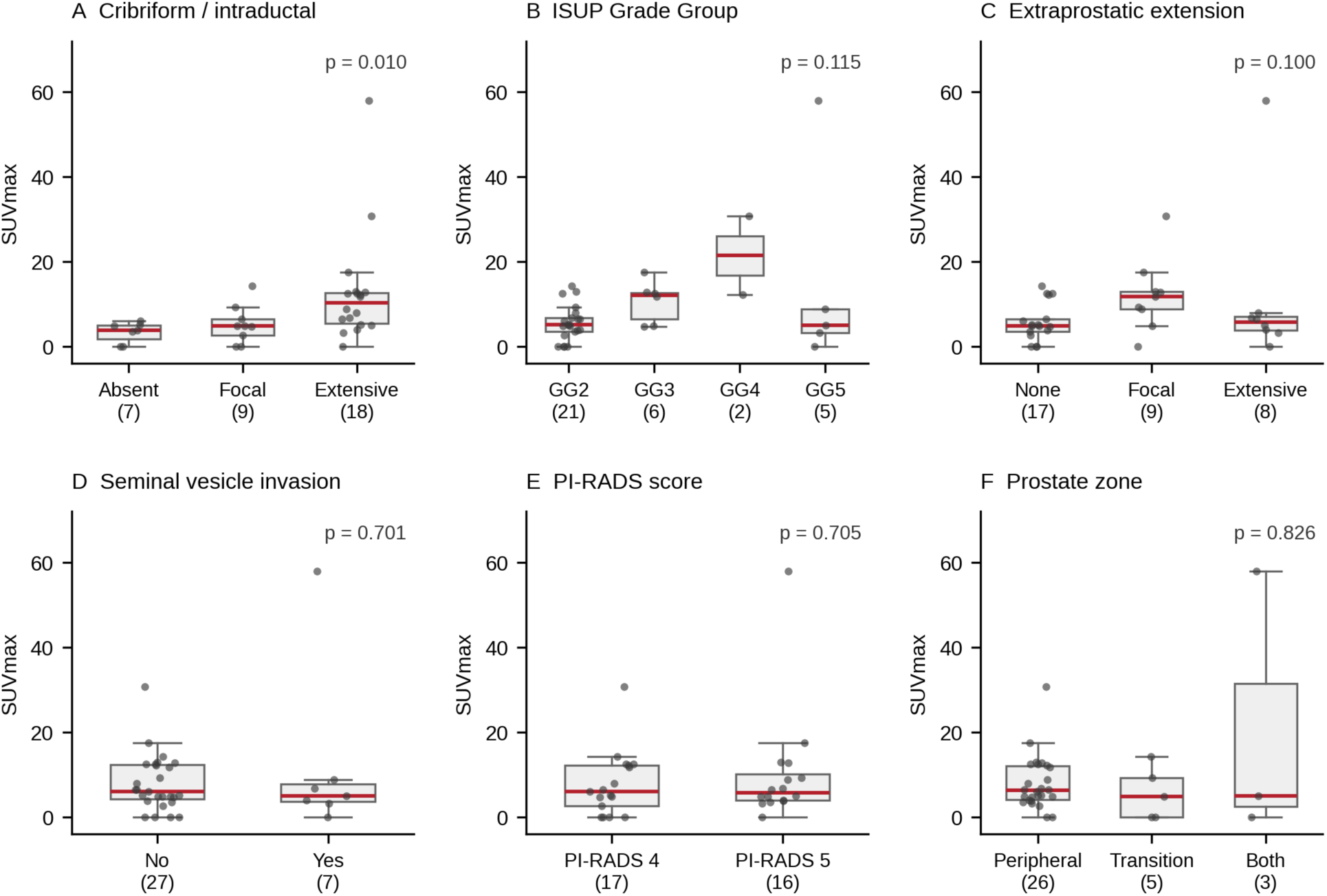
SUVmax across categorical pathologic and imaging features. Box-and-whisker plots of ^18^F-DCFPyL PSMA PET SUVmax for each tumor focus (n = 34), by cribriform/intraductal carcinoma extent (A), ISUP Grade Group (B), extraprostatic extension (C), seminal vesicle invasion (D), PI-RADS score among the 33 MRI-visible foci (E) and prostate zone (F). Boxes show the median and interquartile range, whiskers extend to 1.5× the interquartile range, and individual foci are overlaid as points. Group sizes are given in parentheses on the x axis. p values are from the Kruskal–Wallis test, or the Mann–Whitney test for two-level comparisons, and are unadjusted for multiple comparisons.

**Table 2.** Associations between clinicopathologic, imaging and clinical variables and focus-level SUVmax.

| Variable | Test and result | p value | n |
| --- | --- | --- | --- |
| **Pathology** |  |  |  |
| Grade Group (2 / 3 / 4 / 5) | Kruskal–Wallis; medians 5.1 vs 12.1 vs 21.4 vs 5.0 | 0.115 | 34 |
| % Gleason pattern 4 | $\rho = +0.47$ | 0.005 | 34 |
| % Gleason pattern 5 | $\rho = +0.03$ | 0.880 | 34 |
| Crib/IDC (absent / focal / extensive) | Kruskal–Wallis; medians 3.9 vs 4.8 vs 10.2 | 0.010 | 34 |
| Extraprostatic extension (none / focal / extensive) | Kruskal–Wallis; medians 4.9 vs 11.7 vs 5.8 | 0.100 | 34 |
| Seminal vesicle invasion (no / yes) | Mann–Whitney; medians 6.1 vs 5.0 | 0.701 | 34 |
| Pathologic nodal status (pN0 / pN1) | Mann–Whitney; medians 5.2 vs 12.5 | 0.197 | 34 |
| **Microenvironment** |  |  |  |
| Stromal % | $\rho = -0.40$ | 0.020 | 34 |
| Immune cell % | $\rho = +0.20$ | 0.246 | 34 |
| Stromal category | Kruskal–Wallis; medians 13.4 vs 6.5 vs 4.8 | 0.094 | 34 |
| Immune category | Kruskal–Wallis; medians 4.8 vs 4.7 vs 6.6 | 0.327 | 34 |
| **Tumor size** |  |  |  |
| Greatest dimension, mm | $\rho = +0.26$ | 0.132 | 34 |
| Estimated tumor volume, cm<sup>3</sup> | $\rho = +0.14$ | 0.435 | 34 |
| **MRI** |  |  |  |
| PI-RADS (4 vs 5) | Mann–Whitney; medians 6.1 vs 5.8 | 0.705 | 33 |
| Prostate zone | Kruskal–Wallis; medians 6.3 vs 4.9 vs 5.0 | 0.826 | 34 |
| Standard ADC | $\rho = -0.27$ | 0.130 | 33 |
| Standard DWI signal intensity | $\rho = +0.10$ | 0.578 | 33 |
| T2W signal intensity | $\rho = +0.03$ | 0.856 | 33 |
| High-resolution ADC | $\rho = -0.31$ | 0.096 | 30 |
| High-resolution DWI signal intensity | $\rho = +0.40$ | 0.029 | 30 |
| **Clinical** |  |  |  |
| Baseline PSA | $\rho = +0.04$ | 0.816 | 34 |
| PSA density | $\rho = -0.02$ | 0.918 | 34 |
| MRI prostate volume | $\rho = +0.07$ | 0.702 | 34 |

Spearman rank correlation for continuous variables; Kruskal–Wallis or Mann–Whitney test for categorical variables, with group medians of SUVmax shown in the order listed.
This analysis is exploratory. No correction for multiple comparisons was applied and no individual p value should be read as confirmatory.
Abbreviations: ADC, apparent diffusion coefficient; Crib/IDC, cribriform and/or intraductal carcinoma; DWI, diffusion-weighted imaging; PI-RADS, Prostate Imaging Reporting and Data System; PSA, prostate-specific antigen; SUVmax, maximum standardized uptake value.

Among the continuous covariates, SUVmax correlated positively with % Gleason pattern 4 (Spearman ρ = +0.47, p = 0.005) and inversely with visually estimated stromal content (ρ = −0.40, p = 0.020). High-resolution DWI signal intensity showed a weaker positive correlation (ρ = +0.40, p = 0.029). Tumor greatest dimension (ρ = +0.26, p = 0.132), immune-cell percentage (ρ = +0.20, p = 0.246), standard ADC (ρ = −0.27, p = 0.130), high-resolution ADC (ρ = −0.31, p = 0.096) and T2-weighted signal intensity (ρ = +0.03, p = 0.856) were not associated with SUVmax, nor were other clinical indices (**Figure 2**, **Table 2**).

**Figure 2.**
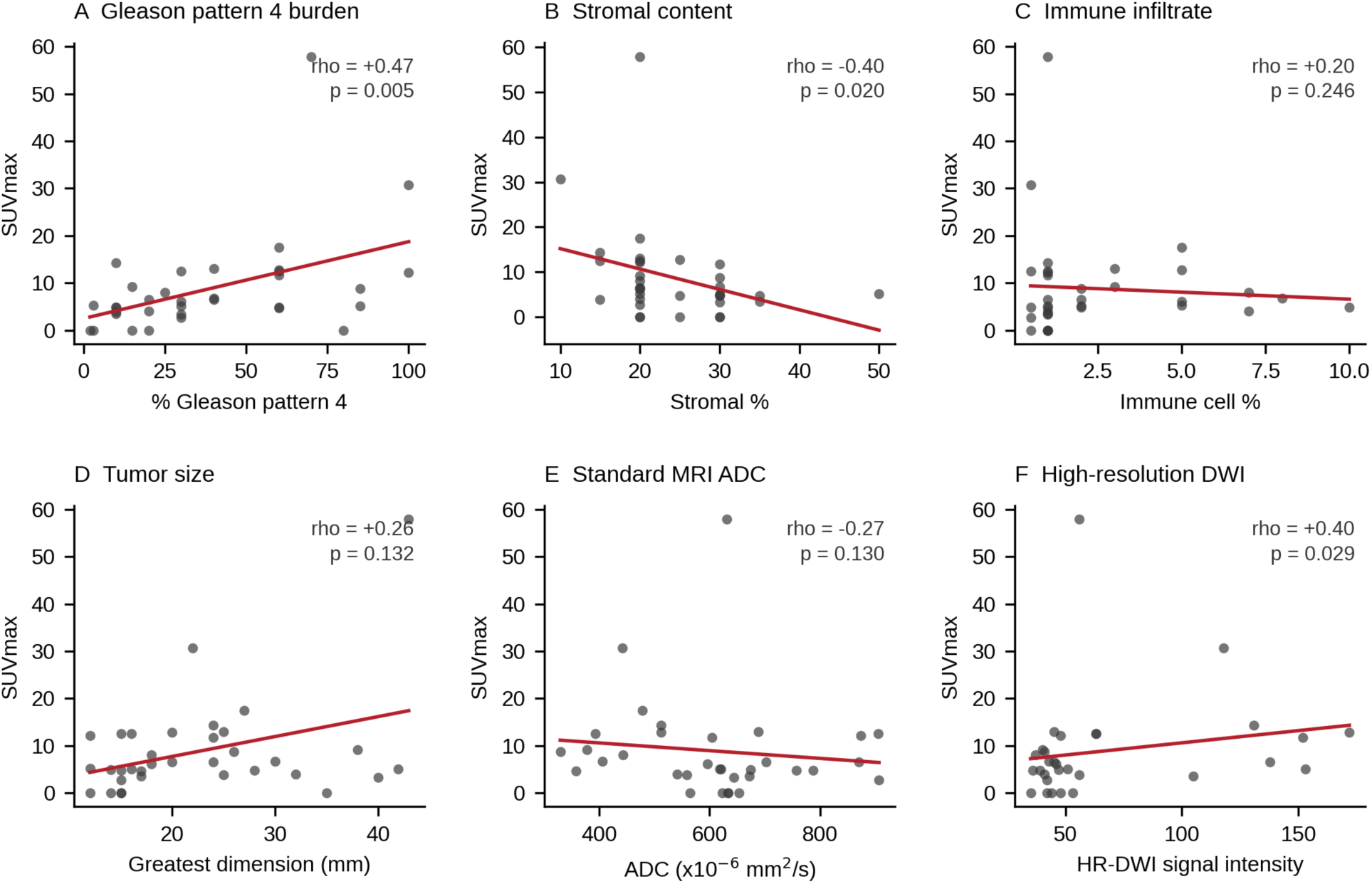
SUVmax against continuous pathologic, microenvironmental and imaging covariates. Scatter plots of tumor-focus SUVmax against % Gleason pattern 4 (A), visually estimated stromal percentage (B), visually estimated immune-cell percentage (C), greatest pathologic dimension (D), standard mpMRI ADC (E) and high-resolution DWI signal intensity (F). Red lines are ordinary least-squares fits shown to indicate direction only. ρ is the Spearman rank-correlation coefficient with its unadjusted p value. n = 30–34 foci per panel depending on covariate availability.

### Adjusted analyses

In separate linear models of log(1+SUVmax), each adjusted for GG and tumor greatest dimension and standardized to 1 SD, % Gleason pattern 4 was associated with higher SUVmax (standardized β = +0.76, 95% CI +0.11 to +1.41, p = 0.024). Crib/IDC architecture (β = +0.36, 95% CI −0.03 to +0.74, p = 0.069) and stromal content (β = −0.30, 95% CI −0.62 to +0.02, p = 0.068) showed concordant trends of similar magnitude, and immune-cell percentage a weaker positive estimate (β = +0.18, 95% CI −0.17 to +0.53, p = 0.296). No MRI metric was associated with SUVmax after adjustment (**Figure 3**).

**Figure 3.**
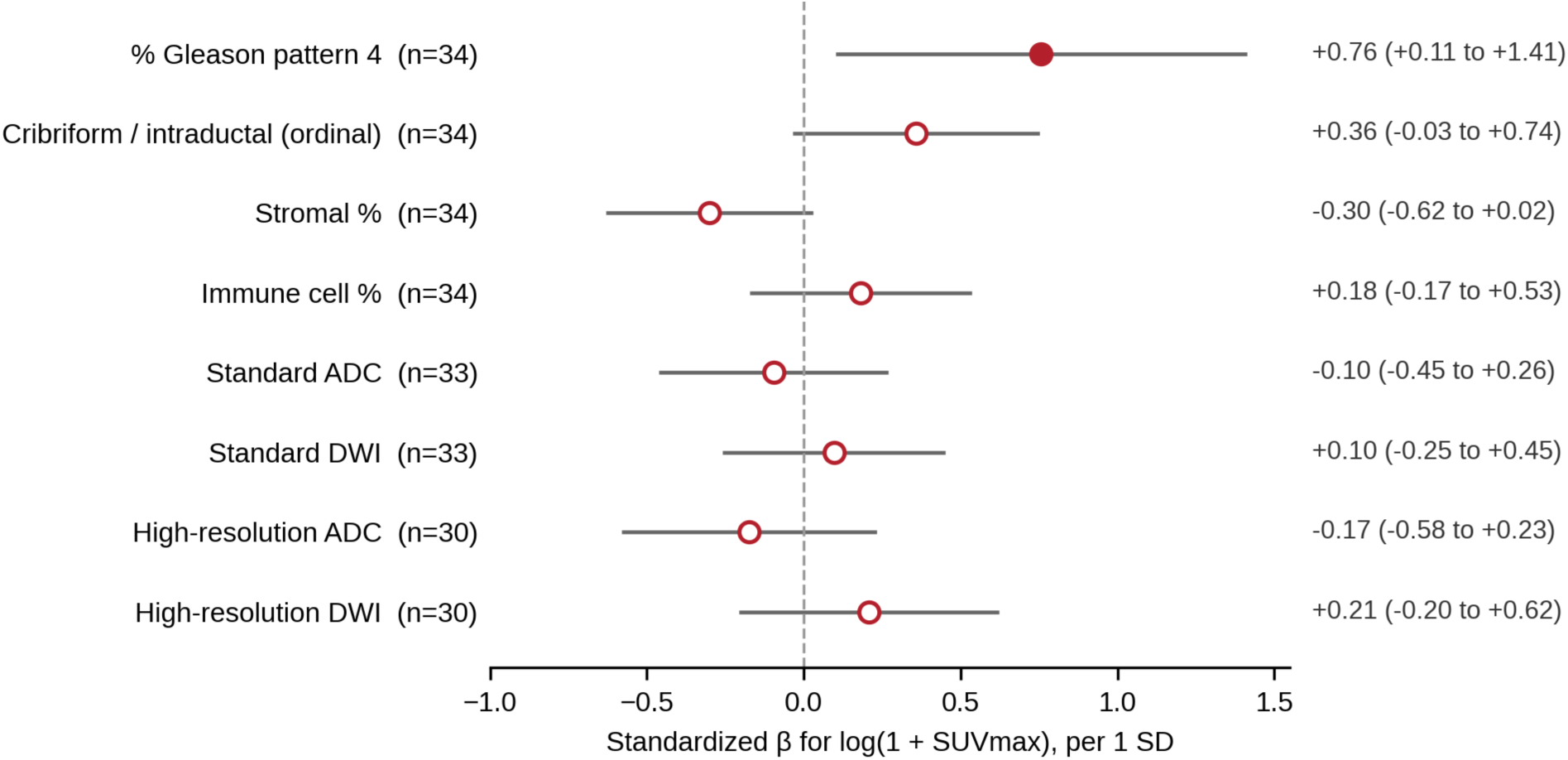
Adjusted associations with intraprostatic PSMA SUVmax. Each row is a separate ordinary least-squares regression of log(1+SUVmax) on the indicated predictor, standardized to 1 SD and adjusted for ISUP Grade Group (ordinal factor: GG2, GG3, GG4–5) and tumor greatest dimension. Markers are standardized β coefficients with 95% confidence intervals; filled markers indicate p < 0.05. Because Grade Group is defined in part by the proportion of Gleason pattern 4, the pattern 4 coefficient reflects within-grade variation in pattern 4 burden.

### Digital pathology re-quantification

A trained pixel classifier was applied to 18 H&E slides from 17 patients that did not contribute training swatches. For immune content the intraclass correlation with pathologist visual estimate was 0.438 (95% CI −0.038 to 0.748) and the rank correlation between methods was ρ = +0.27 (p = 0.28). Mean values were similar across the cohort, with a Bland–Altman mean difference of −0.17 percentage points, but the 95% limits of agreement spanned −4.87 to +4.54 percentage points, a range several times the median immune percentage in this cohort, so agreement at the level of an individual slide was poor (**Figure 4A**). Neither pre-specified concordance criterion was therefore met. For stroma (intraclass correlation 0.048, 95% CI −0.150 to 0.332) and tumor (0.038, −0.152 to 0.318) the disagreement was both in rank and in absolute value: QuPath stroma was on average 22.7 percentage points higher and QuPath tumor 22.6 percentage points lower than the visual estimate (**Figures 4B and 4C; Figure S4**). Rank correlations between the two methods were also near zero for stroma (ρ = +0.14) and tumor (ρ = +0.05).

**Figure 4.**
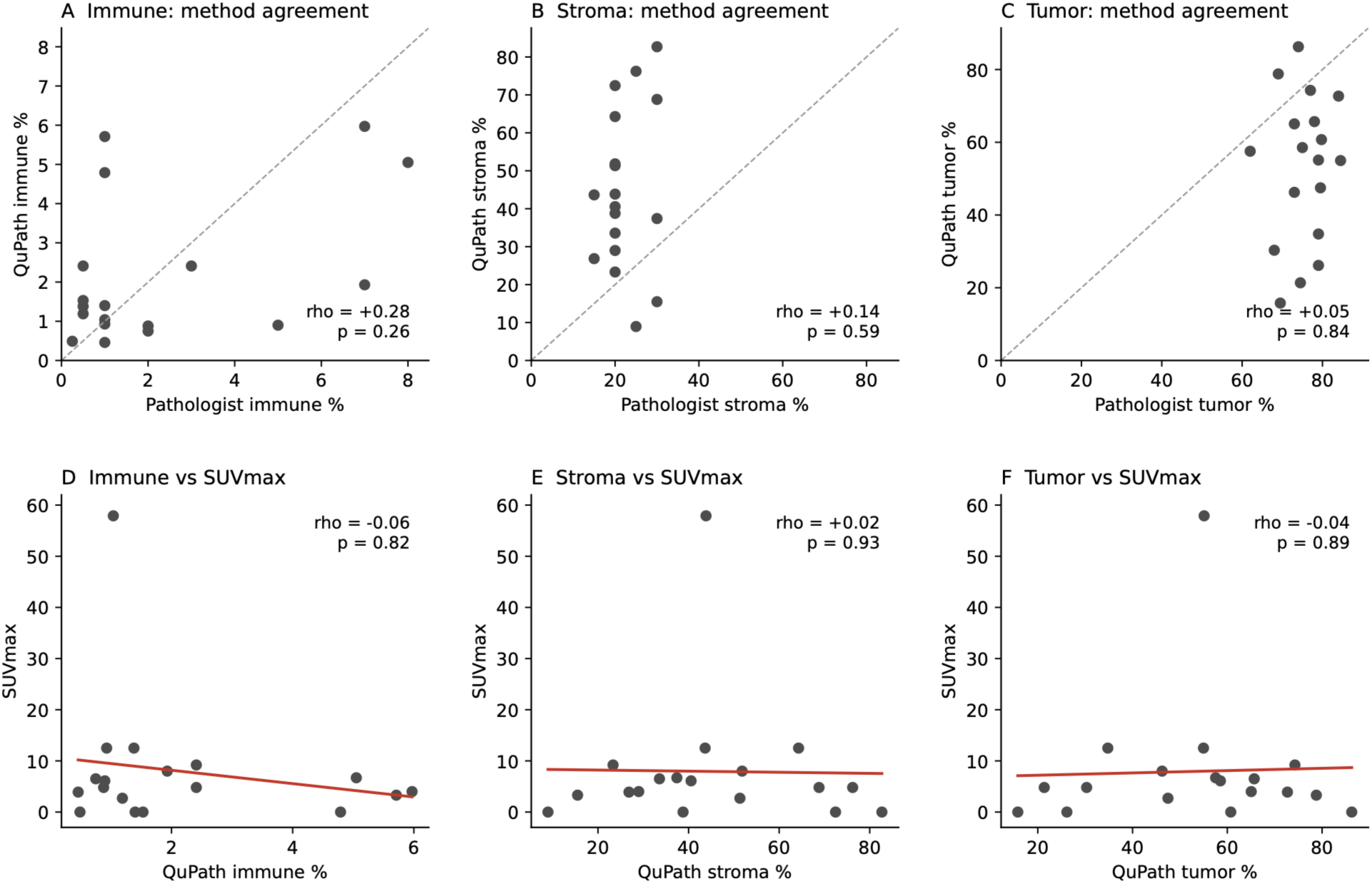
Digital-pathology re-quantification of the tumor microenvironment. A QuPath pixel classifier was applied to 18 H&E slides from 17 patients. These sections are separate from those on which the pathologist visual estimates were made, although cut from the same tumor blocks. Top row: QuPath pixel-area fraction against the pathologist visual estimate for immune (A), stromal (B) and tumor (C) compartments; the dashed line is the line of identity. Bottom row: QuPath compartment fraction against SUVmax for immune (D), stromal (E) and tumor (F) compartments, with ordinary least-squares fits in red. ρ is the Spearman rank-correlation coefficient with its unadjusted p value. Abbreviations: ADC, apparent diffusion coefficient; Crib/IDC, cribriform and/or intraductal carcinoma; DWI, diffusion-weighted imaging; GG, ISUP Grade Group; PI-RADS, Prostate Imaging Reporting and Data System; PSMA, prostate-specific membrane antigen; SD, standard deviation; SUVmax, maximum standardized uptake value.

None of the pixel-based compartment fractions was associated with SUVmax: immune ρ = −0.06 (p = 0.81), stroma ρ = −0.02 (p = 0.94) and tumor ρ = −0.04 (p = 0.89) (**Figures 4D–4F**). The pixel-based re-quantification therefore did not reproduce the visually estimated microenvironment associations described above.

### Foci with low or absent uptake

Nine of the 34 foci (26.5%) had SUVmax below the pre-specified threshold of 4, and five of these had no measurable uptake above background. All nine were MRI-visible. Two were GG5 foci carrying extensive Crib/IDC, extensive EPE and SVI: a 40-mm focus with 30% Gleason pattern 4 and SUVmax 3.3, and a 35-mm focus with 80% Gleason pattern 4 and no measurable uptake. Both were PI-RADS 5. A full comparison of foci above and below the threshold is given in **Supplemental Results** (**Table S4, Figure S5**).

## Discussion

Intraprostatic PSMA SUVmax varied across tumors and tumor characteristics in this well annotated prospective cohort. Cribriform and intraductal architecture, Gleason pattern 4 burden and stromal content each associated with SUVmax uptake, whereas Grade Group, MRI metrics, baseline PSA and PSA density did not. The association between % Gleason pattern 4 and SUVmax was the only one seen in both the univariable screen and the covariate-adjusted model. In this context of limited sample size, heterogeneity of SUVmax appears to be independent of most clinical, pathologic, and MRI features.

PSMA PET metrics do not appear to be a restatement of standard risk factors including GG, PSA and stage. Prior work from our group has linked PSMA expression in treatment-naïve disease to androgen-receptor signaling and treatment susceptibility.^8^ The most parsimonious interpretation is that SUVmax is a readout of a biological axis, plausibly related to differentiation state and androgen-receptor output, that other features do not reflect. This justifies ongoing efforts to characterize the biology that underlies SUVmax heterogeneity.

The burden of Gleason pattern 4 was the only factor that was associated with SUVmax after multivariable adjustment. Mounting evidence has shown that % Gleason pattern 4 has a strong association with oncologic outcomes for localized prostate cancer.^14^ Similar positive trends were noted with Crib/IDC architecture which has also been associated with adverse outcomes.^15^ The inverse, albeit univariable association with stromal content fits expectation, since a focus with more stroma has proportionally less PSMA-expressing epithelium within the same volume of interest, and stromal-infiltration signatures in prostate cancer mark a distinct transcriptional state.^10^ Therefore, architectural changes in the prostate with relevant cancer phenotype implications appear to be reflected in PSMA SUVmax.

Foci with little or no uptake deserve comment. Two GG5 tumors with extensive cribriform and intraductal carcinoma, extraprostatic extension and seminal vesicle invasion were essentially invisible on PSMA PET, one of them entirely so, and both were PI-RADS 5 on MRI. Low-PSMA phenotypes exist in treatment-naïve disease.^8,16–20^ Mechanistically, dedifferentiation with partial loss of androgen-receptor signaling, neuroendocrine-like features and intratumoral heterogeneity of FOLH1 expression all reduce PSMA surface density independently of proliferative rate or grade.^16,21,22^ For the clinician the practical consequence is that a low SUVmax in a PI-RADS 4 or 5 lesion did not indicate a lower likelihood of biologically aggressive disease.

This study has several limitations. First, 34 foci from 27 patients is a small sample. The analysis had 80% power only for correlations of |ρ| ≥ 0.46 at α = 0.05, so effect estimates are imprecise, the confidence intervals rather than the p values should be read as the primary expression of that uncertainty, and a null result here indicates that an association is not large rather than that none exists. Second, the analysis is exploratory and no correction for multiple comparisons was applied; of 23 univariable tests four reached p < 0.05 against approximately one expected by chance, so individual p values should not be interpreted as confirmatory evidence and every association reported here requires replication. Third, % Gleason pattern 4 and Grade Group overlap by definition and their contributions cannot be fully separated at this sample size. Fourth, cribriform and intraductal architecture were scored together and their extent graded semiquantitatively rather than against a fixed area threshold, so the prevalence of extensive disease reported here is not directly comparable across studies. Fifth, foci with no measurable uptake were recorded as SUVmax = 0, a convention that denotes uptake indistinguishable from benign prostatic background; no patient-specific benign prostatic background SUV was recorded prospectively, so uptake in these foci cannot be quantified further, and residual quantification error at the low end of the SUV range would disproportionately affect them. Sixth, immune and stromal percentages were estimated visually and were not reproduced by pixel-classifier re-quantification.

## Conclusions

In this prospective, lesion-level radiology–pathology cohort, intraprostatic PSMA PET SUVmax varied with Crib/IDC architecture, Gleason pattern 4 burden and stromal content but not other indices. PSMA PET metrics appear to be additive to conventional risk factors rather than a restatement of them. Individual high-grade foci with adverse architecture nonetheless showed little or no uptake, so a low SUVmax should not be read as evidence against aggressive disease. Ongoing genomic evaluations based on PSMA PET metrics are warranted to assess the biology that underlies variations in SUVmax and how SUVmax might be levaged to predict biology and personalize cancer care.

## Work Performed At

Departments of Urology, Imaging, and Pathology and Laboratory Medicine, Cedars-Sinai Medical Center, Los Angeles, CA.

## Clinical Trial Registration

ClinicalTrials.go identifier NCT04461509.

## Ethics Approval Statement

The parent trial and this post hoc analysis were approved by the Cedars-Sinai Medical Center Institutional Review Board (STUDY00003844).

## Patient Consent Statement

All participants provided written informed consent in accordance with the parent-trial protocol.

## Data Availability Statement

The de-identified, lesion-level source data supporting this study will be provided upon request. The analysis code that reproduces all reported results is openly available at https://github.com/CamilleSimo0602/psma-pet-suvmax-radiopathologic and archived at Zenodo (DOI: 10.5281/zenodo.22182404).

## Permission to Reproduce Material from Other Sources

All figures and tables are original to this work; no previously published material is reproduced.

