## Supplemental Methods for "PSMA PET SUVmax Adds Information Beyond Conventional Risk Indices in Localized Prostate Cancer: A Prospective Lesion-Level Radiopathologic Study"

Motchoffo Simo et al.

**Contents**

Table S1. Imaging acquisition parameters.

Table S2. Secondary PSMA PET metrics, reference-tissue values and orthogonal tumor dimensions.

Table S3. Per-slide QuPath pixel-classifier measurements, pathologist visual estimates and PSMA PET values.

Table S4. Comparison of foci with SUVmax ≥ 4 and SUVmax < 4.

Figure S1. Study flow diagram.

Figure S2. Prostate cylindrical coordinate system.

Figure S3. Representative QuPath pixel-classifier overlay.

Figure S4. Bland–Altman analysis of QuPath versus pathologist estimates.

Figure S5. Comparison of foci with SUVmax ≥ 4 and SUVmax < 4.

Supplemental Methods.

Supplemental Results.

**Supplemental Methods**

**Parent trial design and eligibility**

Eligibility criteria for the parent trial (NCT04461509) were men ≥18 years with biopsy-proven prostate adenocarcinoma scheduled for either radical prostatectomy (RP) or focal high-intensity focused ultrasound. Exclusion criteria included prior treatment for prostate cancer (surgery, radiation, cryotherapy or high-intensity focused ultrasound), prior androgen-deprivation therapy, contraindication to MRI, contraindication to gadolinium-based contrast, body weight exceeding the scanner bore limit, and inability to provide informed consent. Only patients who underwent RP with specimens available for central prostatectomy pathology review contributed to the present secondary analysis.

**PET–pathology lesion matching**

Each tumor focus recorded on the central prostatectomy pathology map was matched to a specific volume of interest (VOI) on the PSMA PET and MRI images. Matching was performed by a genitourinary pathologist (D.J.L.) using the prostate cylindrical coordinate system (Figure S2) to identify the corresponding anatomic region on the co-registered PET/MRI volume. The VOI was drawn on the fused PET/MRI image and cross-checked against the high-resolution MRI for anatomic correspondence. This per-focus re-quantification necessarily required knowledge of the pathology tumor map, so the imaging reviewers were not blinded to pathology at the matching step. SUVmax is a maximum-voxel metric and is therefore robust to moderate VOI-boundary variability, whereas volume-based metrics are more sensitive to VOI delineation and should be interpreted accordingly. Ambiguous matches, particularly for small multifocal disease and lesions near the capsule, were reviewed jointly with a nuclear-medicine investigator (A.D.). The research PET re-quantification step was added after initial review of the parent-trial reads revealed that clinical reads had been performed at the whole-prostate rather than the per-focus level.

**Prostatectomy pathology annotation protocol**

Specimens were fixed in 10% neutral buffered formalin for a minimum of 24 hours, weighed, inked in four quadrants with distinct colors, and sectioned at approximately 3 mm intervals perpendicular to the urethra from apex to base. For prostates <30 g all sections were submitted, following ISUP submission guidelines. For prostates ≥30 g a partial-submission protocol was applied in which every other section was initially processed, any grossly visible tumor was submitted in its entirety, and any residual grossly visible lesion was submitted at a second pass. All sections carried ordered anatomic designations to permit three-dimensional reconstruction. Cribriform architecture was defined per the 2019 ISUP consensus as a confluent cribriform pattern with a continuous lumen spanning ≥50% of a gland cross-section.

**Tumor volume estimation**

Tumor volume was estimated from three orthogonal pathologic dimensions using the ellipsoid formula, V = (π/6) × w × h × l. The ellipsoid approximation was chosen rather than planimetry because dimensional measurements were available for all foci and are standard in routine prostatectomy reporting, and because the ellipsoid estimate is the pathology analogue of the Prostate Imaging Reporting and Data System (PI-RADS) recommended lesion volume estimate. It systematically overestimates the volume of irregularly shaped foci; this limitation applies uniformly across the cohort.

**Quantitative MRI metrics and ADC non-interchangeability**

For each lesion the following were recorded from standard mpMRI: T2-weighted signal intensity, diffusion-weighted signal intensity at b = 1400 s/mm², apparent diffusion coefficient (ADC, ×10⁻⁶ mm²/s) and three-dimensional lesion dimensions. High-resolution MRI diffusion-weighted signal intensity and ADC were recorded separately. High-resolution ADC values are expected to be lower than standard ADC values for the same lesion, because of reduced partial-volume averaging at the higher spatial resolution (approximately 2.8 versus 15.4 mm³ voxel volume) and because the lower maximum b-value (600 versus 1400 s/mm²) affects the monoexponential fit differently. The two are therefore not interchangeable and are analyzed as separate covariates throughout.

**PET quantification**

For each pathology-confirmed focus a VOI was placed on the PSMA PET images using syngo.via (Siemens Healthineers). PSMA volume was defined by a 50%-of-SUVmax isocontour. Metrics recorded were SUVmax (maximum single-voxel value within the VOI; the primary outcome, selected because it is the most widely reported PSMA PET parameter and is less sensitive to operator-dependent VOI delineation than volume-based metrics), SUVmean, PSMA volume, PSMA-total (SUVmean × PSMA volume) and the heterogeneity index (SUVmax/SUVmean, calculable for the 29 foci with SUVmean >0). All SUV values were normalized to body weight. Reference-tissue SUVmax was measured in the left ventricular cavity or descending aorta (blood pool), the L3 vertebral body (bone marrow) and the parotid gland, and was available for all 34 foci; tumor-to-reference ratios were calculated by dividing tumor SUVmax by the respective reference value.

**Technical considerations**

MR-based attenuation correction on hybrid PET/MRI systems is a recognized source of SUV quantification error, particularly near air–tissue interfaces in the pelvis. Both attenuation-corrected and non-attenuation-corrected images were reconstructed. Partial-volume effects were not corrected for and are acknowledged as a source of SUV underestimation in smaller lesions; foci with a greatest dimension <10 mm were excluded from the analytic cohort to mitigate this. The use of b = 1400 s/mm² for standard diffusion-weighted imaging improves lesion conspicuity but limits comparability of ADC values with institutions using lower maximum b-values.

**Visually estimated microenvironment variables**

Immune cell percentage was estimated visually as the proportion of lymphocytes, macrophages and plasma cells within the tumor area, and was categorized as low (<1%), exactly 1% or high (>1%) based on the observed distribution across foci in this cohort; this threshold is data-driven and exploratory. Five foci were recorded as the literal value <1% and were coded as 0.5 for analysis. Stromal percentage was categorized as low (<20%), moderate (20% to <30%) or high (≥30%). Both categorizations were defined before the adjusted analyses and applied consistently.

**Digital pathology**

Whole-slide H&E images (SVS format, 20× magnification, 0.5037 µm/px) were imported into a QuPath v0.7.0 project and set to brightfield (H&E). For each slide, hematoxylin and eosin stain vectors were estimated using the Visual Stain Editor on a manually selected tumor- and stroma-containing region, with automated exclusion of unrecognized colors; vectors were accepted if the eosin blue component fell within the range observed across the cohort (B = 0.35–0.41). Because that range was derived from the same slides to which it was applied, it functioned as an internal consistency check on stain estimation rather than as an independent quality threshold. For each tumor-focus slide a region of interest was manually annotated to encompass the tumor focus used for pathologic grading and composition scoring. A pixel classifier (Random Trees; Gaussian and weighted-deviation features at σ = 1, 2 and 4 pixels; resolution approximately 2 µm/px) was

trained on annotated swatches covering four classes (tumor, stroma, immune cells, other) distributed across six slides, with at least 20 swatches per class, and was trained iteratively until tumor–stroma borders were visually accurate on held-out slides. Training annotations were reviewed by a genitourinary pathologist (D.J.L.). The classifier was applied to 26 samples that had undergone H&E scanning; five were excluded at quality control for diffuse spread or artifact, leaving 21 quality-passing slides, with three being excluded due to their participation in the training swatches (832-001-B8, 832-008-D16, 832-010-D16). The full analyzed set included 18 slides from 17 distinct patients, one of whom contributed two slides. Per-slide composition was expressed as the percentage of analyzable region-of-interest area occupied by each class. A representative overlay is shown in Figure S3.

Agreement with pathologist visual estimates was assessed by intraclass correlation (ICC; two-way, absolute agreement, single measures) and Bland–Altman analysis, with Spearman rank correlation as a secondary concordance statistic. An ICC of at least 0.60 for immune percentage was pre-specified as the primary concordance criterion and a Spearman rank correlation of at least 0.50 as the secondary criterion; neither was met. A systematic offset in absolute values was anticipated for the stroma and tumor compartments, because pathologist estimates approximate the fraction of the field occupied by tumor-containing areas including intervening stroma whereas the classifier measures pixel-area fractions of malignant epithelium alone. An offset of that kind shifts absolute values without altering the order in which slides rank, so it does not account for disagreement in rank; rank agreement was therefore assessed separately from absolute agreement, and the two are reported separately in the main text. Because one patient contributed two of the 18 slides, no clustering adjustment was applied, given the single duplicated cluster and the exploratory nature of this analysis.

**Supplemental Results**

**Secondary PSMA PET metrics**

Descriptive values for the secondary PET metrics are given in Table S2. These were not part of the analyses reported in the main text and are described here for completeness. SUVmean tracked the cribriform/intraductal (Crib/IDC) ordinal gradient (Spearman ρ = +0.53, p = 0.001) but not Grade Group (ρ = +0.26, p = 0.136). PSMA volume was unrelated to Crib/IDC (ρ = +0.01, p = 0.947), as was PSMA-total (ρ = +0.28, p = 0.115) and the heterogeneity index (ρ = +0.06, p = 0.756; n = 29 foci with SUVmean >0). The tumor-to-blood-pool ratio showed the same gradient across Crib/IDC as SUVmax itself (ρ = +0.56, p < 0.001), and the tumor-to-bone-marrow ratio a weaker one (ρ = +0.33, p = 0.057). Because the tumor-to-reference ratios are derived from SUVmax they are not independent of the primary outcome and were not modeled.

**Foci with SUVmax below the pre-specified threshold**

Nine of 34 foci (26.5%) had SUVmax < 4 and five had no measurable uptake above background (recorded SUVmax = 0). Median SUVmax was 0.0 in this group versus 8.0 in the 25 foci at or above the threshold. All nine were MRI-visible, comprising 5 of 17 PI-RADS 4 lesions (29%) and 4 of 16 PI-RADS 5 lesions (25%). Seven of the nine were GG2 and two were GG5. The five foci with no measurable uptake ranged from 12 to 35 mm in greatest dimension; the largest, a 35-mm GG5 focus, is well above the size at which partial-volume effects alone would be expected to suppress SUVmax to background.

Adverse histologic features were on the whole more common in foci at or above the threshold, consistent with the cohort-level association between Crib/IDC and uptake reported in the main text: Crib/IDC was absent in 4 of 9 foci below the threshold (44%) versus 3 of 25 at or above it (12%), and extraprostatic extension was present in 3 of 9 (33%) versus 14 of 25 (56%). Seminal vesicle invasion was evenly distributed (2 of 9 versus 5 of 25). Tumor size, stromal percentage, standard ADC and baseline PSA did not differ between the groups (Table S4, Figure S5). Visually estimated immune-cell percentage was lower below the threshold (median 1.0% versus 2.0%, p = 0.014), a difference that was not reproduced by pixel-classifier re-quantification on the 18 re-scored slides, in which median immune percentage was 1.40% below the threshold and 1.38% at or above it (p = 0.98; main text, Figure 4). This comparison did not survive an independent method of measurement and is reported as hypothesis-generating only.

Two foci met the pre-specified clinical-mismatch definition of GG ≥3 with SUVmax < 4, both GG5, both with extensive Crib/IDC, extensive extraprostatic extension and seminal vesicle invasion, both PI-RADS 5 and both node-negative. The first was a 40-mm focus with 30% Gleason pattern 4, SUVmax 3.3, SUVmean 2.0 and standard ADC 644 ×10⁻⁶ mm²/s; its tumor-to-blood-pool ratio of 6.6 is inflated by the lowest blood-pool reference value in the cohort (0.5) and should not be read as high uptake. The second was a 35-mm focus with 80% Gleason pattern 4 and no measurable uptake (SUVmax 0.0, SUVmean 0.0, standard ADC 654 ×10⁻⁶ mm²/s). With two events this outcome cannot be modeled and is described here only.

**Anatomic location and multifocality**

No location or multifocality variable was associated with SUVmax. Base involvement carried the strongest signal: median SUVmax 6.6 in the 22 foci involving the base versus 4.4 in the 12 that did not (p = 0.060). Apex involvement was null (25 versus 9 foci; p = 0.907), as were anteroposterior position (p = 0.161; posterior n = 27, anterior n = 4, both n = 1, unspecified n = 2, the last retained as a separate level rather than excluded) and laterality (p = 0.617; right n = 17, left n = 12, bilateral n = 5). Mid-gland involvement was present in all 34 foci and was therefore not testable. Multifocality, defined at the patient level as multifocal carcinoma on the central pathology map, was also null: 22 foci from multifocal patients (median SUVmax 5.7) versus 12 from patients with unifocal disease (5.9; p = 0.745). Seven patients contributed two analyzed foci each; within-patient correlation on the modeled log scale was negligible (intraclass correlation <0.01).

**Table S1. Imaging acquisition parameters.**

| **Parameter** | **Value** |
| --- | --- |
| **Standard mpMRI: T2-weighted** |  |
| Sequence | 2D turbo spin-echo |
| Slice thickness | 3.5 mm |
| In-plane resolution | 2.1 × 2.1 mm² |
| **Standard mpMRI: diffusion-weighted** |  |
| Sequence | Single-shot echo-planar imaging |
| b-values | 0, 100, 400, 1400 s/mm² |
| Voxel volume | ~15.4 mm³ |
| ADC units | ×10⁻⁶ mm²/s |
| **Standard mpMRI: dynamic contrast-enhanced** |  |
| Contrast agent | Gadobutrol |
| Dose | 0.1 mmol/kg |
| Injection rate | 2 mL/s |
| Temporal resolution | 5 s |
| Phases | 50 |
| **High-resolution MRI** |  |
| Sequence | Diffusion-prepared bSSFP, 3D multishot readout |
| In-plane resolution | 0.9 × 0.9 mm² |
| Slice thickness | 3.5 mm |
| Voxel volume | ~2.8 mm³ |
| Field of view | 180 × 180 mm² |
| Matrix | 208 × 208 |
| TR/TE | 1,200/80 ms |
| b-values | 0, 300, 600 s/mm² |
| Bandwidth | 801 Hz/pixel |
| Scan time | 7 min 32 s |
| **PSMA PET** |  |
| Radiotracer | ¹⁸F-DCFPyL (PyL; Lantheus) |
| Injected activity | 333 MBq (9 mCi, ±20%) |
| Uptake time | 60 min post-injection |
| Acquisition duration | ~30 min |
| Field of view | Skull base to mid-thigh (most); pelvis-only (subset) |
| Mode | 3D |
| Time per bed position | 3 min |
| Reconstruction | OSEM with point-spread-function correction |
| Iterations / subsets | 3 / 21 |
| Attenuation correction | MR-based (Dixon) |
| Output | Attenuation-corrected and non-attenuation-corrected images |

All imaging was performed on a Siemens Biograph mMR hybrid PET/MRI scanner (Siemens Healthineers, Erlangen, Germany) in a single session. Abbreviations: ADC, apparent diffusion coefficient; bSSFP, balanced steady-state free precession; OSEM, ordered-subsets expectation maximization; TE, echo time; TR, repetition time.

**Table S2. Secondary PSMA PET metrics, reference-tissue values and orthogonal tumor dimensions.**

| **Metric** | **Overall (n = 34 foci)** |
| --- | --- |
| **Orthogonal tumor dimensions** |  |
| Craniocaudal, mm | 18.0 [12.8, 26.8] |
| Anteroposterior, mm | 16.5 [12.5, 22.8] |
| Transverse, mm | 8.0 [6.2, 10.8] |
| **Secondary PET metrics** |  |
| SUVmean | 3.5 [2.4, 6.8] |
| PSMA volume, cm³ | 0.9 [0.6, 2.8] |
| PSMA-total | 5.1 [2.6, 9.7] |
| Heterogeneity index (n = 29) | 1.68 [1.60, 1.78] |
| **Reference-tissue SUVmax and ratios** |  |
| Blood pool | 1.45 [1.30, 1.80] |
| Bone marrow | 1.00 [0.80, 1.35] |
| Parotid gland | 22.20 [14.20, 25.00] |
| Tumor-to-blood-pool ratio | 3.95 [2.50, 7.62] |
| Tumor-to-bone-marrow ratio | 4.76 [3.29, 12.75] |

Values are median [IQR]. The heterogeneity index (SUVmax/SUVmean) is calculable only where SUVmean >0, which held for 29 of 34 foci. Reference-tissue values were available for all 34 foci.

**Table S3. Per-slide QuPath pixel-classifier measurements, pathologist visual estimates and PSMA PET values (n = 18 slides from 17 patients).**

| **Slide** | **QuPath** |  |  | **Pathologist** |  |  | **SUVmax** | **SUVmean** |
| --- | --- | --- | --- | --- | --- | --- | --- | --- |
|  | **Immune** | **Stroma** | **Tumor** | **Immune** | **Stroma** | **Tumor** |  |  |
| 832-020-D8 | 0.90 | 40.55 | 58.54 | 5.00 | 20.00 | 75.00 | 6.1 | 3.40 |
| 832-022-B14 | 5.71 | 15.49 | 78.79 | 1.00 | 30.00 | 69.00 | 3.3 | 2.00 |
| 832-023-C7 | 4.79 | 8.95 | 86.26 | 1.00 | 25.00 | 74.00 | 0.0 | 0.00 |
| 832-024-B21 | 0.46 | 26.84 | 72.69 | 1.00 | 15.00 | 84.00 | 3.9 | 2.40 |
| 832-029-B4 | 0.93 | 64.29 | 34.78 | 1.00 | 20.00 | 79.00 | 12.5 | 6.80 |
| 832-029-B7 | 1.40 | 72.45 | 26.14 | 1.00 | 20.00 | 79.00 | 0.0 | 0.00 |
| 832-034-E5 | 2.41 | 23.32 | 74.27 | 3.00 | 20.00 | 77.00 | 9.2 | 5.70 |
| 832-036-D6 | 1.04 | 43.85 | 55.11 | 1.00 | 20.00 | 79.00 | 57.9 | 13.00 |
| 832-040-D20 | 1.19 | 51.34 | 47.47 | 0.50 | 20.00 | 79.50 | 2.7 | 1.50 |
| 832-041-D23 | 0.49 | 38.78 | 60.73 | 0.25 | 20.00 | 79.75 | 0.0 | 0.00 |
| 832-042-E4 | 1.53 | 82.69 | 15.78 | 0.50 | 30.00 | 69.50 | 0.0 | 0.00 |
| 832-044-D3 | 2.41 | 76.23 | 21.36 | 0.50 | 25.00 | 74.50 | 4.8 | 2.70 |
| 832-045-D6 | 5.97 | 28.99 | 65.05 | 7.00 | 20.00 | 73.00 | 4.0 | 2.50 |
| 832-046-D12 | 1.93 | 51.83 | 46.24 | 7.00 | 20.00 | 73.00 | 8.0 | 5.10 |
| 832-049-D18 | 5.05 | 37.41 | 57.53 | 8.00 | 30.00 | 62.00 | 6.7 | 3.90 |
| 832-050-D20 | 0.75 | 33.57 | 65.68 | 2.00 | 20.00 | 78.00 | 6.5 | 3.60 |
| 832-051-F16 | 0.88 | 68.81 | 30.31 | 2.00 | 30.00 | 68.00 | 4.8 | 2.70 |
| 832-052-D7 | 1.38 | 43.65 | 54.97 | 0.50 | 15.00 | 84.50 | 12.5 | 6.60 |

All values are percentages of the analyzable region of interest except SUVmax and SUVmean. Slides are labeled by subject and block. Subject 832-029 contributed two slides.

**Table S4. Comparison of foci with SUVmax ≥ 4 and SUVmax < 4.**

| **Characteristic** | **SUVmax ≥ 4 (n = 25)** | **SUVmax < 4 (n = 9)** |
| --- | --- | --- |
| **PSMA PET metrics** |  |  |
| SUVmax | 8.0 [5.1, 12.5] | 0.0 [0.0, 3.3] |
| SUVmean | 4.8 [3.1, 7.5] | 0.0 [0.0, 2.0] |
| PSMA volume, cm³ | 1.1 [0.7, 3.6] | 0.0 [0.0, 1.4] |
| PSMA-total | 7.5 [3.7, 12.0] | 0.0 [0.0, 2.5] |
| Tumor-to-blood-pool ratio | 5.75 [3.38, 8.67] | 0.00 [0.00, 2.25] |
| Tumor-to-bone-marrow ratio | 7.62 [4.64, 13.00] | 0.00 [0.00, 2.70] |
| **Pathologic features** |  |  |
| Greatest dimension, mm | 22.0 [16.0, 27.0] | 15.0 [15.0, 25.0] |
| Grade Group 2 | 14 (56.0%) | 7 (77.8%) |
| Grade Group 3 | 6 (24.0%) | 0 (0.0%) |
| Grade Group 4 | 2 (8.0%) | 0 (0.0%) |
| Grade Group 5 | 3 (12.0%) | 2 (22.2%) |
| Crib/IDC extensive | 16 (64.0%) | 2 (22.2%) |
| Crib/IDC focal | 6 (24.0%) | 3 (33.3%) |
| Crib/IDC absent | 3 (12.0%) | 4 (44.4%) |
| Extraprostatic extension present | 14 (56.0%) | 3 (33.3%) |
| Seminal vesicle invasion | 5 (20.0%) | 2 (22.2%) |
| **Tumor microenvironment** |  |  |
| Immune cell % | 2.0 [1.0, 5.0] | 1.0 [1.0, 1.0] |
| Stromal % | 20.0 [20.0, 30.0] | 25.0 [20.0, 30.0] |
| **MRI and clinical** |  |  |
| Standard ADC, ×10⁻⁶ mm²/s | 600 [444, 692] | 634 [623, 654] |
| Baseline PSA, ng/mL | 6.9 [6.3, 8.8] | 5.9 [4.0, 10.2] |

Continuous variables are median [IQR]; categorical variables are n (%). The SUVmax threshold of 4 was pre-specified in the parent-trial protocol. Immune cell percentage with <1% coded as 0.5. Standard ADC was available for 33 of 34 foci.

**Figure S1. Study flow diagram.**

Flow of participants through the parent trial (NCT04461509) and into the present lesion-level analysis. Of 62 patients enrolled, 33 were allocated to the radical prostatectomy arm; six were excluded because no tumor focus met the eligibility criteria of Grade Group ≥2 and greatest dimension ≥10 mm on central pathology review, yielding 27 patients contributing 34 tumor foci. Abbreviations: GG, ISUP Grade Group; hrMRI, high-resolution MRI; mpMRI, multiparametric MRI; RP, radical prostatectomy.

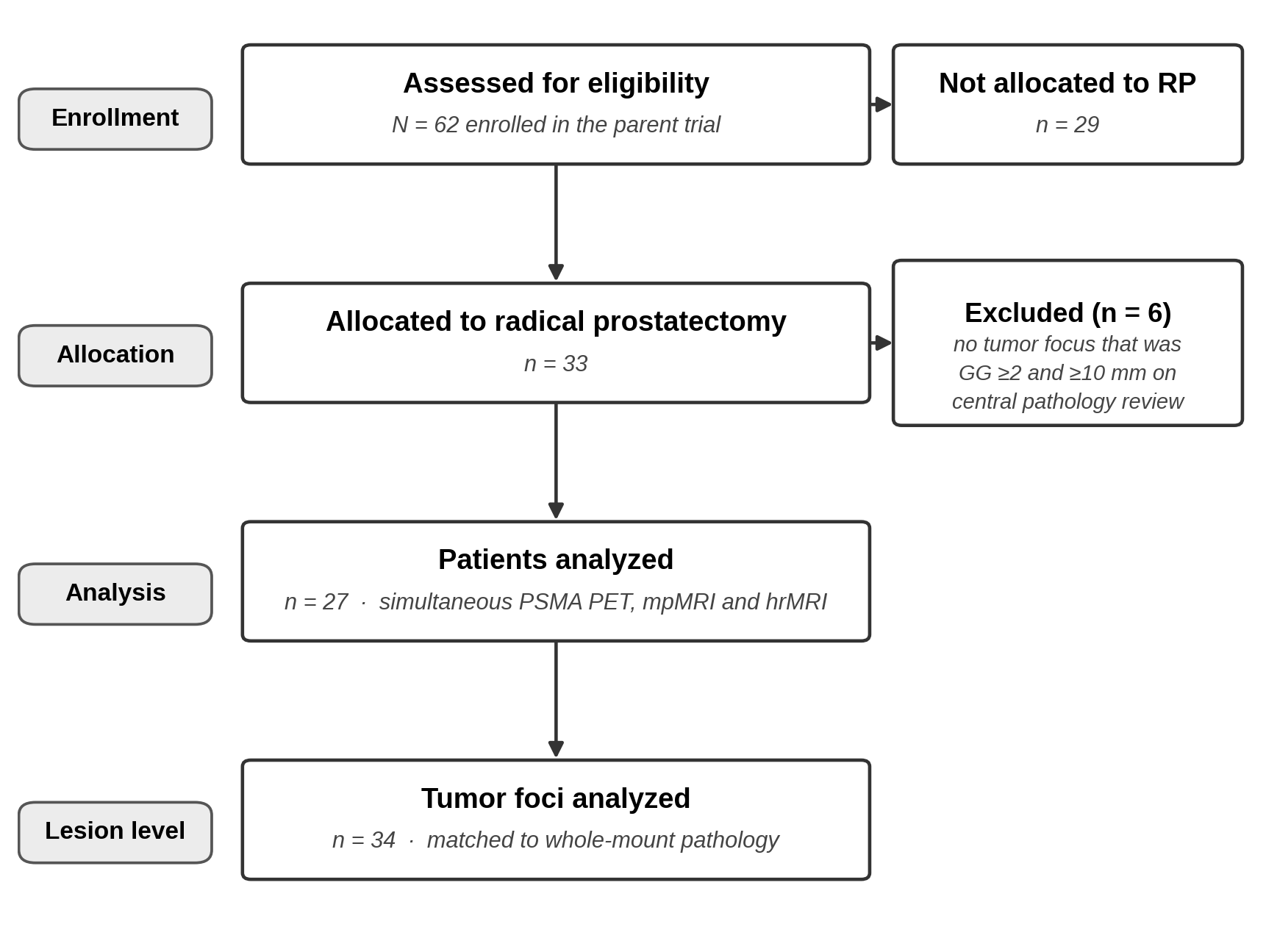

**Figure S2. Prostate cylindrical coordinate system.**

Schematic of the cylindrical coordinate system used to localize tumor foci on surgical histology and to align each pathology-defined focus with its corresponding lesion on PSMA PET and MRI. The prostate is treated as a right-circular cylinder with the urethra on the long axis; each focus is annotated by axial level (apex, mid, base), circumferential clock-face sector and radial zone (peripheral, transition or central). The same frame is applied to the PET/MRI volume of interest during research re-quantification, allowing one-to-one matching between the pathology tumor map and the imaging focus.

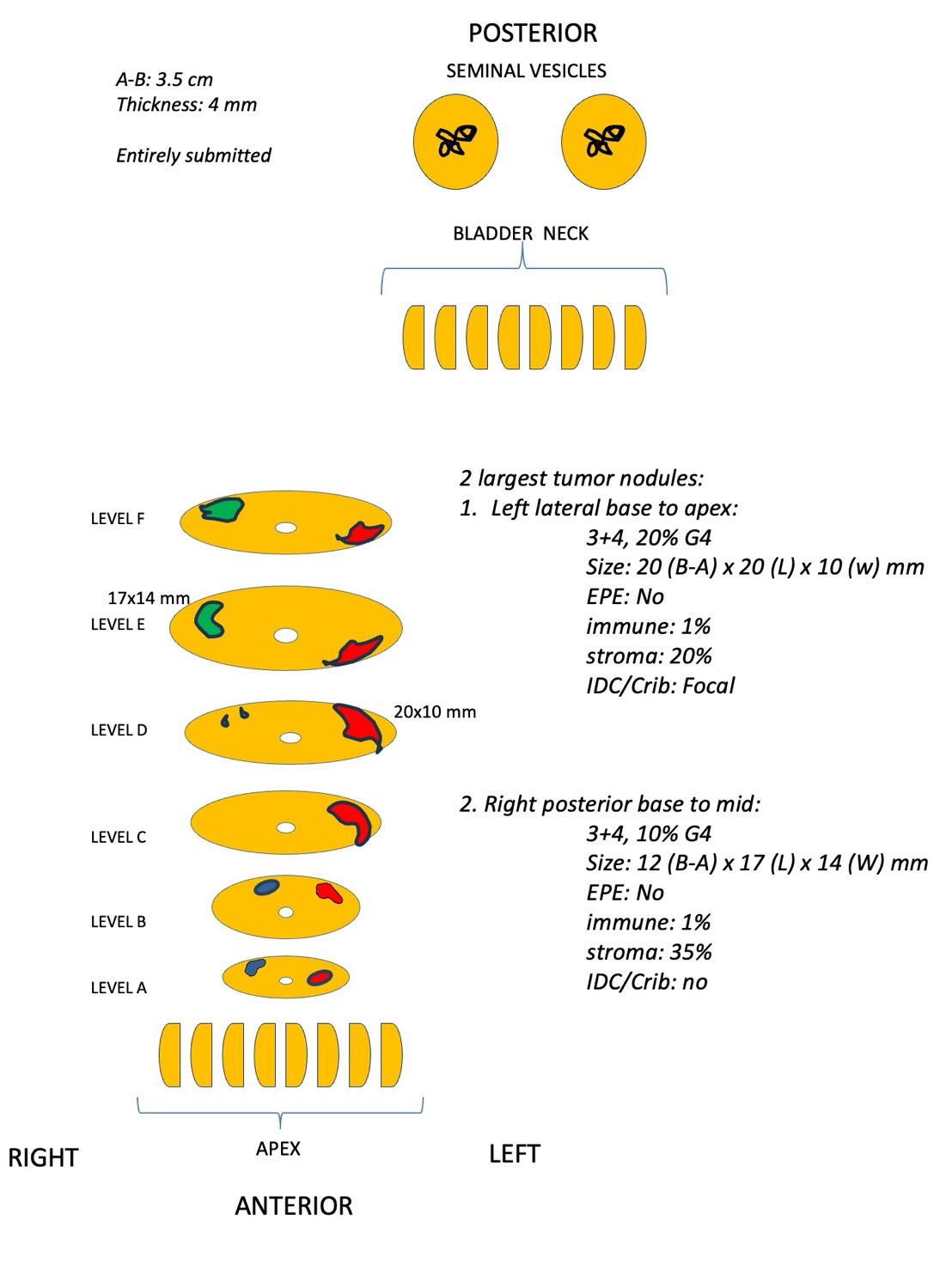

**Figure S3. Representative QuPath pixel-classifier overlay.**

Trained pixel-classifier overlay on a representative H&E section. Tumor is shown in red, stroma in green and immune cells in purple.

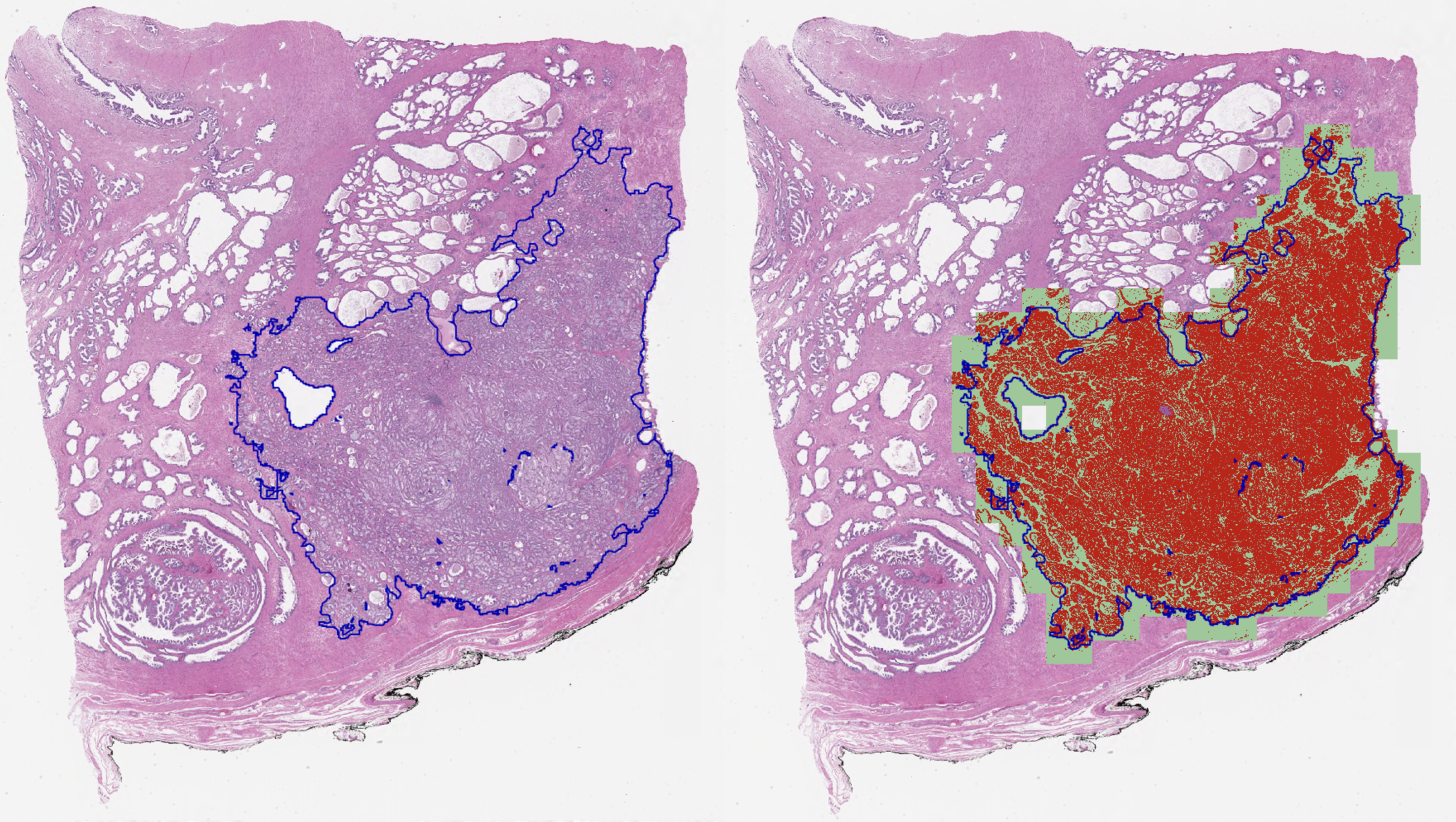

**Figure S4. Bland–Altman analysis of QuPath versus pathologist estimates.**

Difference between the QuPath pixel-area fraction and the pathologist visual estimate, plotted against the mean of the two, for the immune, stromal and tumor compartments across 18 slides. The solid line is the mean difference and the dashed lines the 95% limits of agreement. The two measurements were made on different physical sections from the same tumor block, so section-to-section heterogeneity contributes to the observed limits of agreement. The large offsets for stroma and tumor reflect a definitional difference between the two methods rather than measurement error. A definitional offset alone would leave the rank ordering of slides intact, however, and the ordering also disagreed, so the two methods are not interchangeable at the level of an individual slide.

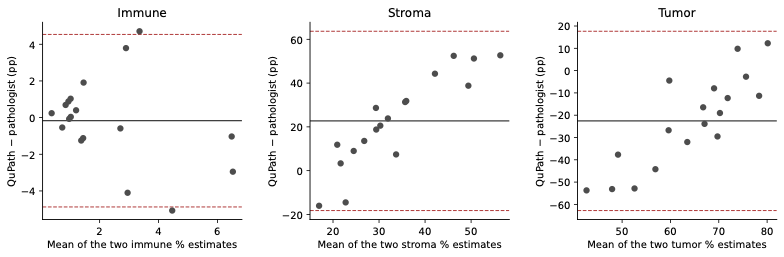

**Figure S5. Comparison of foci with SUVmax ≥ 4 and SUVmax < 4.**

Box-and-whisker plots of greatest pathologic dimension, % Gleason pattern 4, visually estimated stromal percentage and visually estimated immune-cell percentage in the 25 foci with SUVmax ≥ 4 and the 9 foci with SUVmax < 4. Boxes show the median and interquartile range, whiskers extend to 1.5× the interquartile range, and individual foci are overlaid as points. p values are from the Mann–Whitney test and are unadjusted for multiple comparisons.

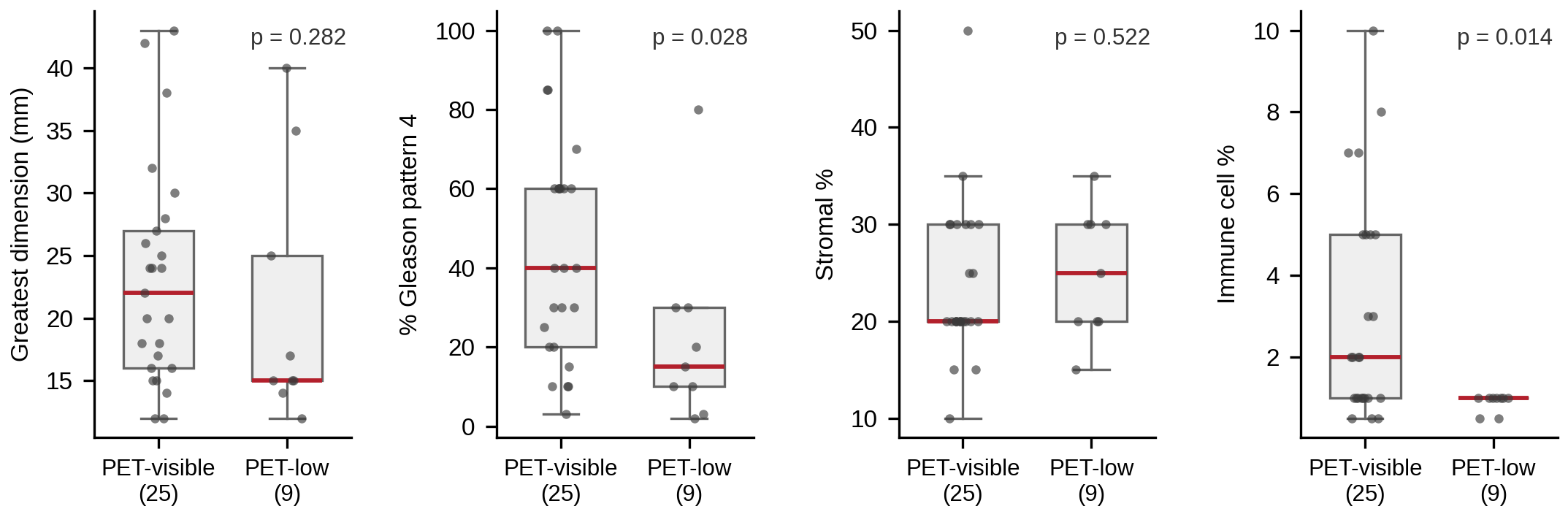
